# A Logistic Formula in Biology and Its Application to Deaths by the Third Wave of COVID-19 in Japan

**DOI:** 10.1101/2021.01.30.21250827

**Authors:** Akira Kokado, Takesi Saito

## Abstract

A logistic formulation in biology is applied to analyze deaths by the third wave of COVID-19 in Japan.

## 1 Introduction

In previous papers [1], [2] we have proposed a logistic formulation of infection. The logistic formula is useful in the population problem in biology. It is closely related to the SIR model [3] in the theory of infection. which is powerful to analyze an epidemic about how it spreads and how it ends [4-11]. The SIR model is composed of three equations for S, I and R, where they are numbers for susceptibles, infectives and removed, respectively. Our logistic formula has been driven approximately from this SIR model. This approximate formula, however, has a simple form, so that it is very useful to discuss an epidemic.

In this paper, our policy is to use mainly data of deaths by COVID-19 in Japan, but not so often of cases.

In Sec. 2, we review the derivation of this logistic formula and construct basic equations used later. In Sec. 3, our logistic formula is applied to the 3^*rd*^ wave of COVID-19 in Japan. The final section is devoted to concluding remarks. We attach Appendix for error estimations.

## 2 The logistic formula

In previous works [1], [2] we found a logistic formula, which is an approximate one of the SIR model [3] in the theory of infection. Let us summarize the formula briefly. The third equation of the SIR model is given by

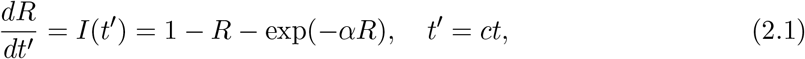

*where R* is the removed number, *I*(*t*′) the infectious number, *α* the basic reproduction number, *c* the removed ratio and *t* the true time. There is another function *S* in the SIR model, which stands for the susceptible number. Three functions *S, I* and *R* are normalized as *S* +*I* +*R* = 1.

Let us expand the exponential factor as, with *x* = *αR*, in the second order,

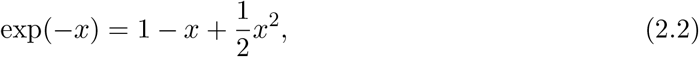

Here we have assumed *x*^2^*/*2 < 1. In this approximation we have

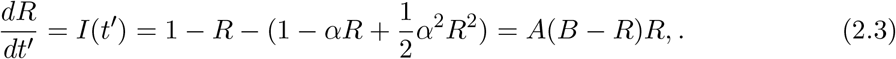

with *A* = *α*^2^*/*2, *B* = 2(*α* − 1)*/α*^2^. This equation is a type of that in the logistic growth curve in Biology, easily solved as

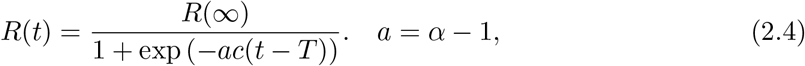

where *AB* = *α* − 1 = *a* and *B* = *R*(∞) = 2(*α* − 1)*/α*^2^ = 2*R*(*T*), *T* being the peak day of infection. Inserting this into Eq.(2.1) we get:

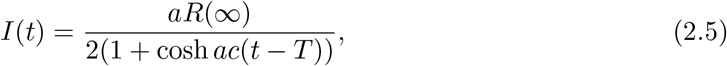

We make use of data [12] of deaths rather than cases. It is our policy not to use data of cases. Define the mortality ratio *λ* by

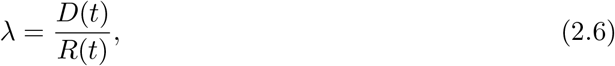

where *D*(*t*) is the accumulated number of deaths. For the population *N*, the total number of deaths at t is given by

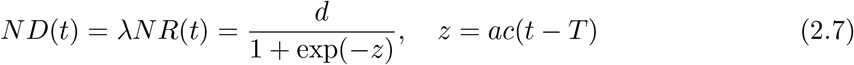

where *d* = *ND*(∞) = *λNR*(∞) stands for the final total number of deaths. Note that from Eq.(2.7) we have a theorem, *ND*(*T*) = *d/*2, which means the total number of deaths at the peak is just a half of the final ones.

Let us rewrite Eq.(2.7) in a form as

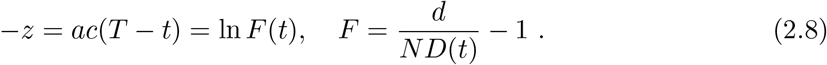

Accordingly, for different times *t*_1_, *t*_2_ and *t*_3_ we have equations

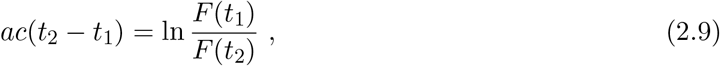

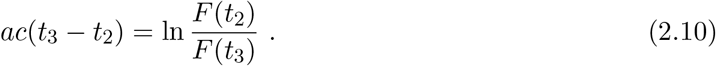

When time differences in Eqs. (2.9) and (2.10) are equal, we get a useful equation for *d*

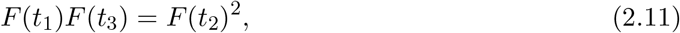

### 3 Application to the third wave of COVID-19

Our logistic formula is applied to the 3^*rd*^ wave of COVID-19 in Japan.

*ND*(*t*) is the accumulated number of deaths in the 3^*rd*^ wave at *t* in Japan, which is an average of deaths for 7 days in a middle at each t with standard deviations, where t is the date starting from Oct. 11. We have subtracted the accumulated number of deaths, 1628, in the 1^*st*^ and 2^*nd*^ waves, from that in the 1^*st*^, 2^*nd*^ and 3^*rd*^ waves.

A use is made of data of deaths [12]. Substituting data on the Table 1 into Eq.(2.11), we get the equation for *d*

**Table 1:** Date and the number of deaths in the 3^*rd*^ wave

| t | N D(t) |
| --- | --- |
| $t_1$=Dec. 17 | $n_1 = 1150.143 \pm 95.106$ |
| $t_2$=Dec. 31 | $n_2 = 1826.714 \pm 106.463$ |
| $t_3$=Jan. 14 | $n_3 = 2673 \pm 105.347$ |

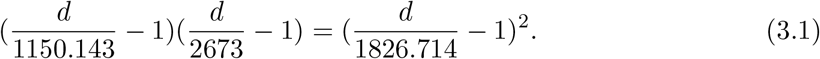

We find a solution of this equation to be *d* = 5810.410. According to the theorem *ND*(*T*) = *d/*2, the number of deaths at the peak *t* = *T* is *d/*2 = 2905.205. The result of *d* = 5810.410 is substituted into Eq.(2.9), then it follows that

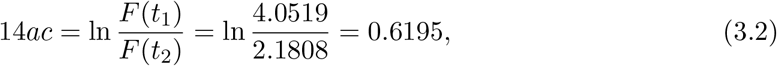

hence

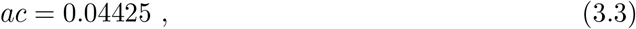

and

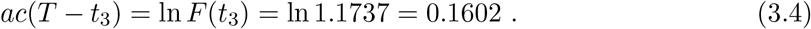

The last equation means *T* − *t*_3_ = 3.62 *≃* 4, hence *T* =Jan.14 + 4 = Jan.18.

Error estimations for *d* = 5810.410 and *T* =Jan.18 (=99) can be seen from Appendix, i.e.,

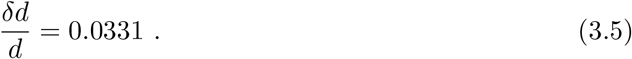

which states *δd* = 192.3 and *δT* = −0.2.

To sum up we have

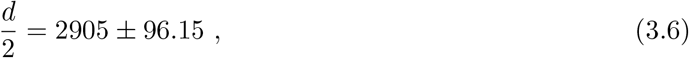

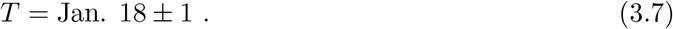

In order to fix a, we make use of *c* = 0.041, which is frequently quoted value. Since *ac* = 0.044, we have *a* = 1.07, that is, *α* = 1 + *a* = 2.07. Once having *α* = 2.07, we can draw curves of *S, I* and *R* by means of Excel in Fig. 1. From this we find *t*′ = 4.86 at the peak of *I*, so that from the formula *t*′ = *ct* we get *c* = *t*′*/T* = 4.86*/*99 = 0.049. This value *c* = 0.049 is slightely different from *c* = 0.041, that is, its error is 16%. About the error we have discussed in a previous paper [1] that the error of our logistic model against the SIR model is about 15%. Therefore, this difference may be allowed.

**Figure 1:**
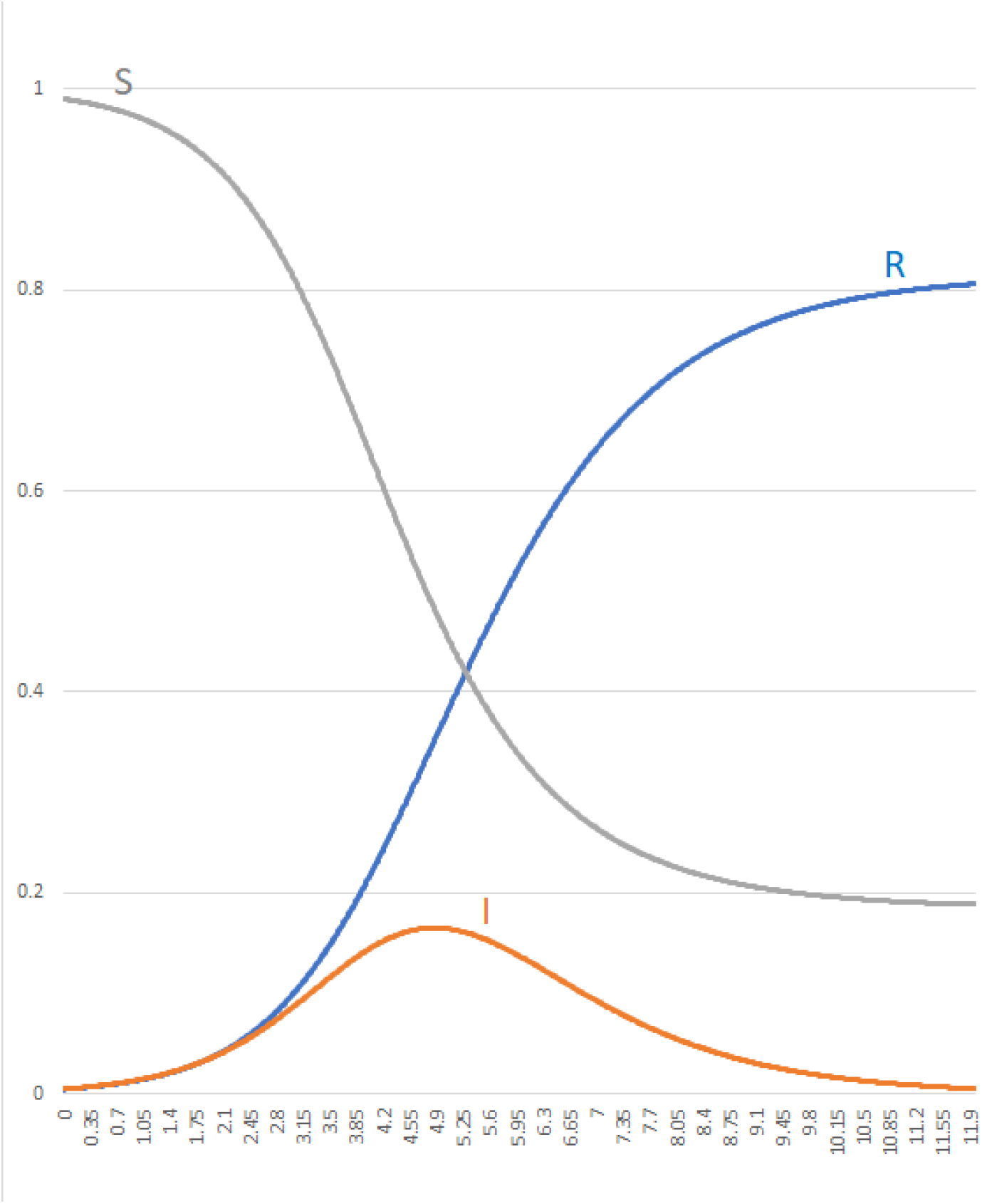
Graph of *S, I* and *R* for *α* = 2.07 and *c* = 0.041

## 4 Concluding remarks

We found that the third wave began from Oct. 11 and will peak at around Jan. 18 *±* 1, 2021 with total deaths 2905 *±* 96. The total number of deaths for the 1^*st*^, 2^*nd*^ and 3^*rd*^ waves on the peak is 2905 *±* 96 + 1628 = 4593 *±* 96. This should be compared with the observed value 4547. The basic reproduction number of the third wave is *α* = 2.07 with the removed ratio *c* = 0.041. Curves of *S, I* and *R* are given in Fig. 1

Let us discuss the population related with infectives. Japanese population is divided into two groups, one is self-isolated and the other is not self-isolated. We can remove the self-isolated group from the infection route, because it is irrelevant to the infection. The non-isolated group is, therefore, relevant to the infection. We put such a population as *N*. From Fig. 1, one can see that

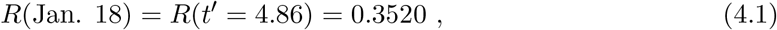

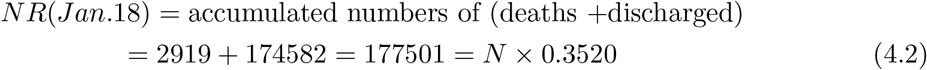

yield *N* = 504264. Therefore, we conclude that the non-isolated population *N* in the 3^*rd*^ wave is about 504,000. We can then estimate the number of infectives on Jan. 18 by formulas

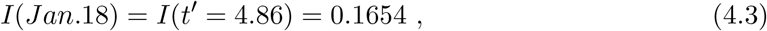

and

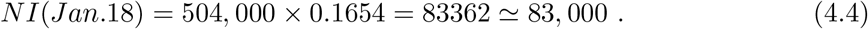

The present number of infectives on Jan. 18 in Japan is about 74,000 and seems to be slightly lower than the calculated value 83,000.

## Data Availability

The availability of all data referred to in the manuscript is OK.

https://toyokeizai.net/sp/visual/tko/covid19

## Acknowledgement

We would like to express our deep gratitude to K. Shigemoto for many valuable discussions and big supports.

## Appendix Error estimations

The total death *d* is given by

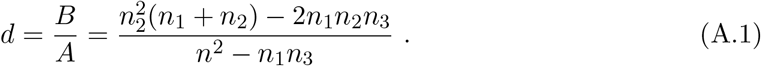

The relative error of *d* is then derived by

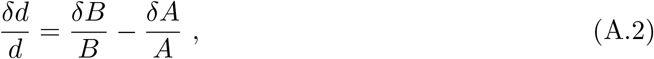

where

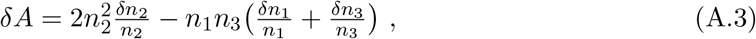

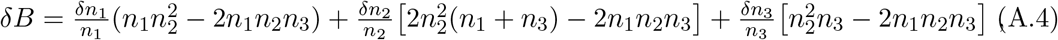

Substituting 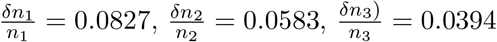 into above, we get

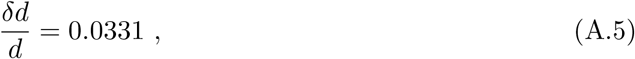

In the same way, from the equation for *ac*

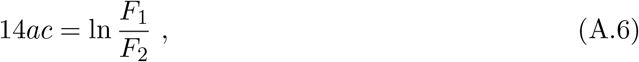

we have

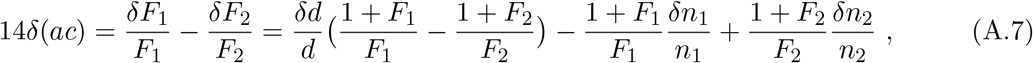

so that

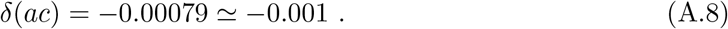

Finally, the equation for the peak day *T, ac*(*T* − *t*_3_) = ln *F*_3_, yields

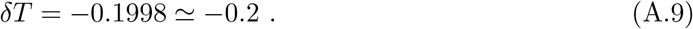

